# The Association Between Alpha-1 Adrenergic Receptor Antagonists and In-Hospital Mortality from COVID-19

**DOI:** 10.1101/2020.12.18.20248346

**Authors:** Liam Rose, Laura Graham, Allison Koenecke, Michael Powell, Ruoxuan Xiong, Zhu Shen, Kenneth W. Kinzler, Chetan Bettegowda, Bert Vogelstein, Susan Athey, Joshua T. Vogelstein, Maximilian F. Konig, Todd H. Wagner

## Abstract

Effective therapies for coronavirus disease 2019 (COVID-19) are urgently needed, and preclinical data suggest alpha-1 adrenergic receptor antagonists (α_1_-AR antagonists) may be effective in reducing mortality related to hyperinflammation independent of etiology. Using a retrospective cohort design with patients in the Department of Veterans Affairs healthcare system, we use doubly robust regression and matching to estimate the association between baseline use of α_1_-AR antagonists and likelihood of death due to COVID-19 during hospitalization. Having an active prescription for any α_1_-AR antagonist (tamsulosin, silodosin, prazosin, terazosin, doxazosin, or alfuzosin) at the time of admission had a significant negative association with in-hospital mortality (relative risk reduction 18%; odds ratio 0.73; 95% CI 0.63 to 0.85; p ≤ 0.001) and death within 28 days of admission (relative risk reduction 17%; odds ratio 0.74; 95% CI 0.65 to 0.84; p ≤ 0.001). In a subset of patients on doxazosin specifically, an inhibitor of all three alpha-1 adrenergic receptors, we observed a relative risk reduction for death of 74% (odds ratio 0.23; 95% CI 0.03 to 0.94; p = 0.028) compared to matched controls not on any α_1_-AR antagonist at the time of admission. These findings suggest that use of α_1_-AR antagonists may reduce mortality in COVID-19, supporting the need for randomized, placebo-controlled clinical trials in patients with early symptomatic infection.

## 1 Introduction

The viral replication phase in Coronavirus disease 2019 (COVID-19) can be followed by a hyperinflammatory host immune response, hereafter referred to as COVID-19-associated hyperinflammation, which can lead to acute respiratory distress syndrome (ARDS), multiorgan dysfunction, and death despite maximal supportive care (1–4). While dexamethasone and other immunosuppressive strategies have shown some promise in improving outcomes in patients with severe COVID-19, they have not shown benefit (and may be detrimental) when given to patients with less advanced disease (5–7). To date, immunomodulatory therapeutic strategies that prevent the development of hyperinflammation and thereby halt progression to severe COVID-19 do not exist.

Catecholamines (adrenaline, noradrenaline, and dopamine) are monoamine hormones that signal through adrenergic receptors (ARs) expressed on tissues including cells of the immune system (8–10). Cells of the innate and adaptive immune system (phagocytes, lymphocytes) are capable of producing catecholamines *de novo* and signal in an autocrine/paracrine self-regulatory fashion (9,11). Beyond their well-established role in neurotransmission and physiological fight-or-flight responses, catecholamines have been shown to amplify immune responses and enhance acute inflammatory injury *in vitro* and *in vivo* by increasing cytokine production in immune cells (e.g., IL-6, TNF-α, MIP-2) (8,10–12). In animal models of hyperinflammation, prophylactic treatment with an alpha-1 adrenergic receptor (α_1_-AR) antagonist that inhibits all three receptor subtypes (α_1A_-, α_1D_-, and α_1B_-AR) can prevent cytokine storm and death by blocking deleterious catecholamine signaling and immune responses (11). In a retrospective analysis of patients hospitalized with acute respiratory distress, patients incidentally taking any α_1_-AR antagonist had a 34% relative risk reduction of being mechanically ventilated and dying (n=16,801, odds ratio 0.70) compared to non-users (13). Similarly, the risk of progression to mechanical ventilation and death was significantly reduced in a retrospective analysis of >300,000 patients hospitalized with pneumonia who were prescribed α_1_-AR antagonists prior to their index admission, suggesting that baseline inhibition of catecholamine signaling may improve clinical outcomes in acute lower respiratory tract infection or inflammation (13). We therefore hypothesized that early treatment with α_1_-AR antagonists can improve mortality and ameliorate disease in patients with symptomatic SARS-CoV-2 infection (14), but data demonstrating the efficacy of α_1_-AR antagonists in COVID-19 specifically is lacking.

The objective of this study was to examine the association of use of α_1_-AR antagonists with in-hospital mortality in patients with COVID-19. Here, we analyzed a large cohort of patients hospitalized at Veterans Health Administration (VA) hospitals, in whom α_1_-AR antagonists are commonly used to treat unrelated diseases such as benign prostatic hyperplasia (BPH), post-traumatic stress disorder (PTSD), or arterial hypertension (15). We hypothesized that patients with COVID-19 taking α_1_-AR antagonists at the time of hospital admission would be less likely to die during their hospitalization.

## 2 Methods

### 2.1 Study Population & Variables

We included all patients admitted to a VA hospital between February 20, 2020, and October 7, 2020 with a confirmed COVID-19 diagnosis (Figure 1). Diagnosis codes for COVID-19 were identified from the Centers for Disease Control and Prevention (CDC) coding guidelines for COVID-19 (16,17). The VA COVID-19 Shared Data Resource was used to identify VA patients with a SARS-CoV-2 laboratory test result (18). This data resource combines VA-specific lab results with non-VA lab results using text extraction from patient medical records. Because over 90% of α_1_-AR antagonist users in the analysis were older men, we excluded women to reduce unmeasured confounding unrelated to COVID-19, specifically with respect to respiratory conditions. We also excluded patients under age 45 and patients over age 85 given the strong relationship between the severity of COVID-19 and age.

**Figure 1.**
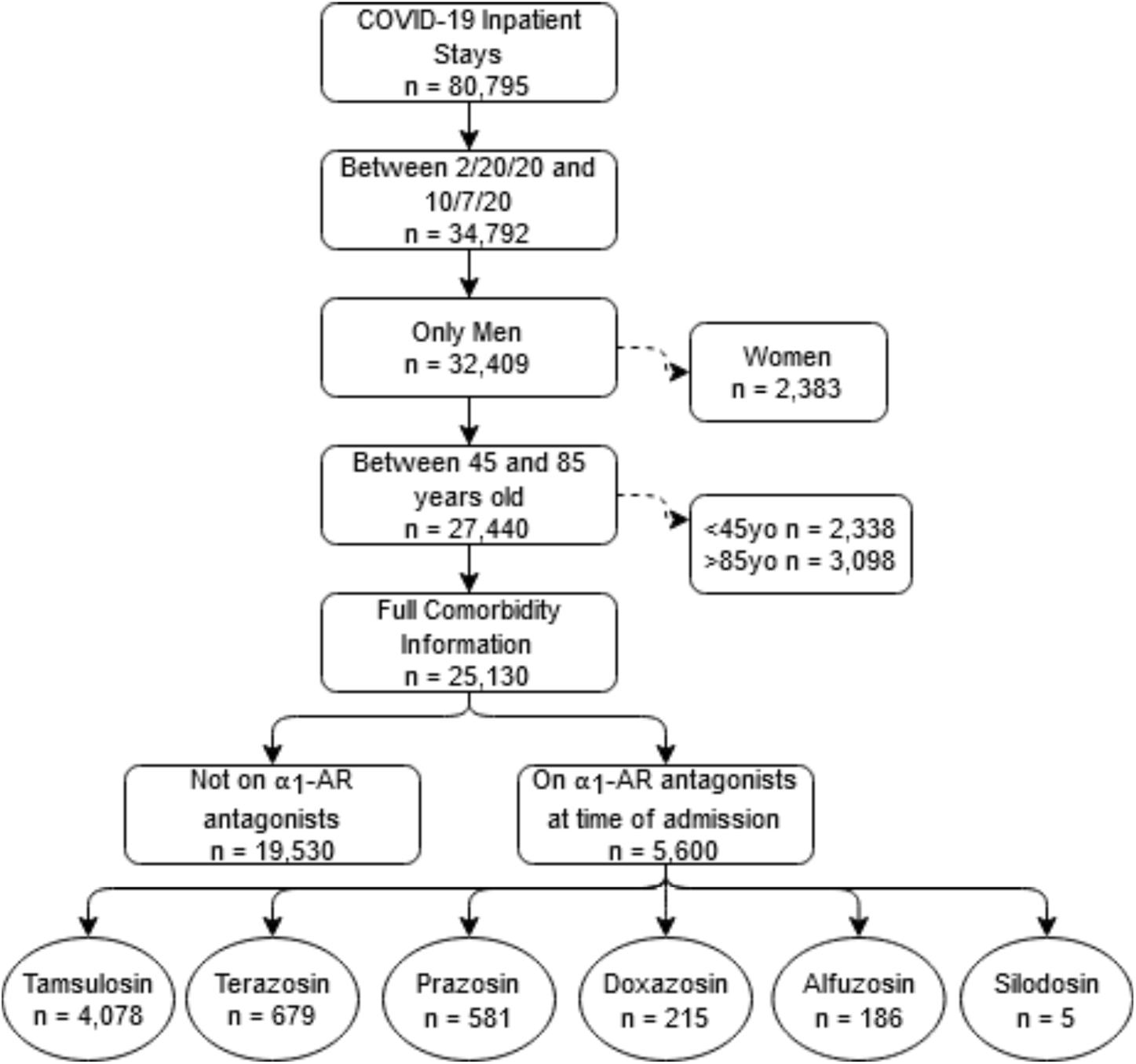
CONSORT Flow Diagram. Consort diagram. Note that the bottom row of medications are not mutually exclusive, with a small number of patients having more than one on hand at time of admission.

An expanded sample included all patients with laboratory-confirmed, “suspected positive”, or “possible positive” COVID-19 according to National COVID Cohort Collaborative (N3C) criteria (19). This Suspected COVID-19 sample excluded patients who tested negative for SARS-CoV-2. To the extent we can measure COVID-19 severity at time of admission, we find that this cohort was not operationally different from the main cohort based on vital signs at time of admission (Supplementary Figure 1).

The primary outcomes were death during the index hospitalization and death within 28 days of admission. The primary exposure variable was the use of α_1_-AR antagonists at the time of admission for the index hospitalization. Active prescriptions of α_1_-AR antagonists (tamsulosin, silodosin, prazosin, doxazosin, alfuzosin, and terazosin) were identified and defined by the patient having medication on hand on the day of the index admission, regardless of dosage. Secondary analyses examined the effect of tamsulosin (the most commonly prescribed α_1_-AR antagonist with selective antagonism on α_1A_- and α_1D_-, but not α_1B_-ARs) and doxazosin (a nonselective antagonist acting on all three α_1_-ARs) individually. Finally, with in-hospital therapies evolving during the pandemic, we repeated the analysis by week and VA hospital to ensure results were not driven by any particular time or location.

We obtained data on patient demographics, vital signs, and prescription drugs from the VA’s corporate data warehouse (CDW). Patient comorbidities were captured based on the International Classification of Diseases, Version 10 codes from VA care in the year prior to index admission. Other physiologic variables, including oxygen saturation and temperature, were defined at time of inpatient admission.

### 2.2 Analysis

Analyses followed the methodology of a companion paper examining patients with acute respiratory distress and pneumonia (13). Unadjusted analysis compared patients with α_1_-AR antagonist prescriptions to all other patients with COVID-19 using Fisher’s exact test. We then estimated propensity scores and trimmed the sample to ensure overlap in the propensity score distributions of the exposed and unexposed groups. On this reduced sample, the adjusted analysis used inverse propensity-weighted logistic regression adjusting for patient age at admission (input as a demeaned cubic polynomial to allow a nonlinear relationship), calendar week, location of hospitalization, and comorbidities diagnosed any time in the two years prior to the index inpatient stay. This approach is “doubly robust” in that it uses the observed confounders in both the calculation of the propensity score and the odds ratios. Comorbidities included in the matching procedure were diabetes mellitus, arterial hypertension, heart failure, ischemic heart disease, acute myocardial infarction, chronic obstructive pulmonary disease (COPD), end-stage renal disease (ESRD), and PTSD. We also included an indicator variable for oxygen saturation under 94 percent on the day of admission.

All of the control variables reflect information on patients prior to admission with COVID-19. As noted above, we controlled for secular changes in COVID-19 care using calendar week, starting with February 20, 2020. We chose not to examine endpoints during the hospital stay, such as use of a ventilator or admission to the ICU, given this is based on physician coding or data structures that we cannot assure were handled uniformly, especially during surges. We also chose not to control for processes of care during the stay given this could introduce bias in the analysis.

We then conducted a 5:1 matched analysis using the same covariates as the adjusted model (10). This approach assigns each exposed patient to a set of five unexposed patients most similar on observed characteristics and does not make assumptions about the functional form of the potential relationship between confounders and the outcome. Matches were selected using a greedy, nearest-neighbor approach based on Mahalanobis distance (11). The matched analysis used the Cochran-Mantel-Haenszel test to obtain odds ratios, confidence intervals, and p-values. We also present relative risk reductions (RRR) for the matched cohorts, and the pre- and post-matching balance of covariates is shown in Figure 3.

**Figure 2:**
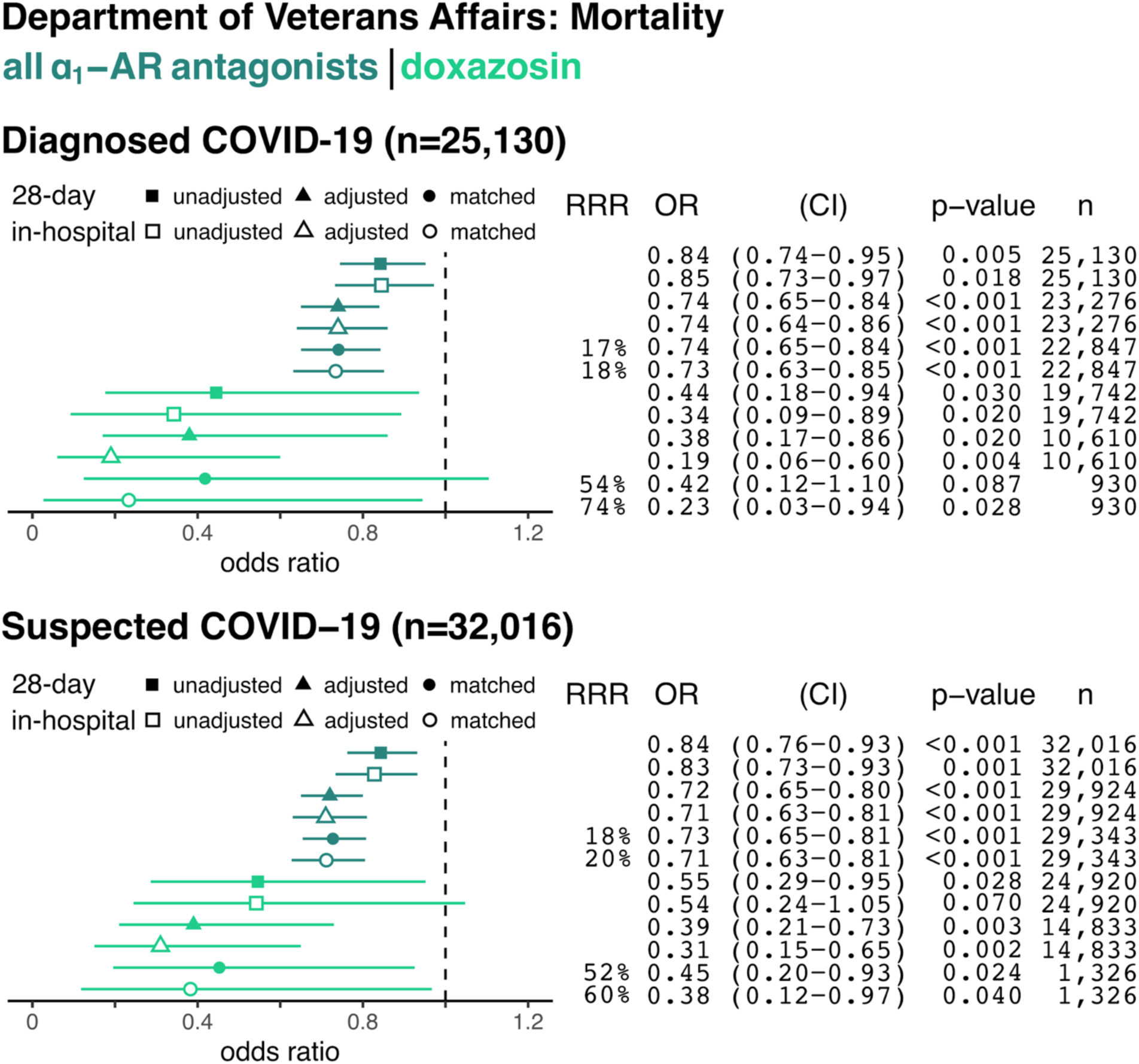
The Association Between Alpha-1 Adrenergic Receptor Antagonists and In-Hospital and 28-Day Mortality from COVID-19. Data are shown for hospitalized patients with confirmed COVID-19 (top panel) and with confirmed plus suspected COVID-19 (e.g., no confirmatory testing available, bottom panel). Forest plots show the odds ratios (OR) for in-hospital mortality based on prior use of any α_1_-AR antagonists (i.e., tamsulosin, silodosin, prazosin, terazosin, doxazosin, or alfuzosin; dark green) or only doxazosin (light green) in each panel. Unadjusted (square), adjusted model (triangle), and matched model (circle) analyses are shown for each sample group. Filled symbols reflect the odds of death within 28 days from index hospital admission (including deaths after discharge), whereas empty symbols reflect odds of death during the index admission. Relative risk reduction (RRR), odds ratios (ORs) for death, 95% confidence intervals (CI), p-values, and sample size (n) for each analysis are shown on the right.

**Figure 3.**
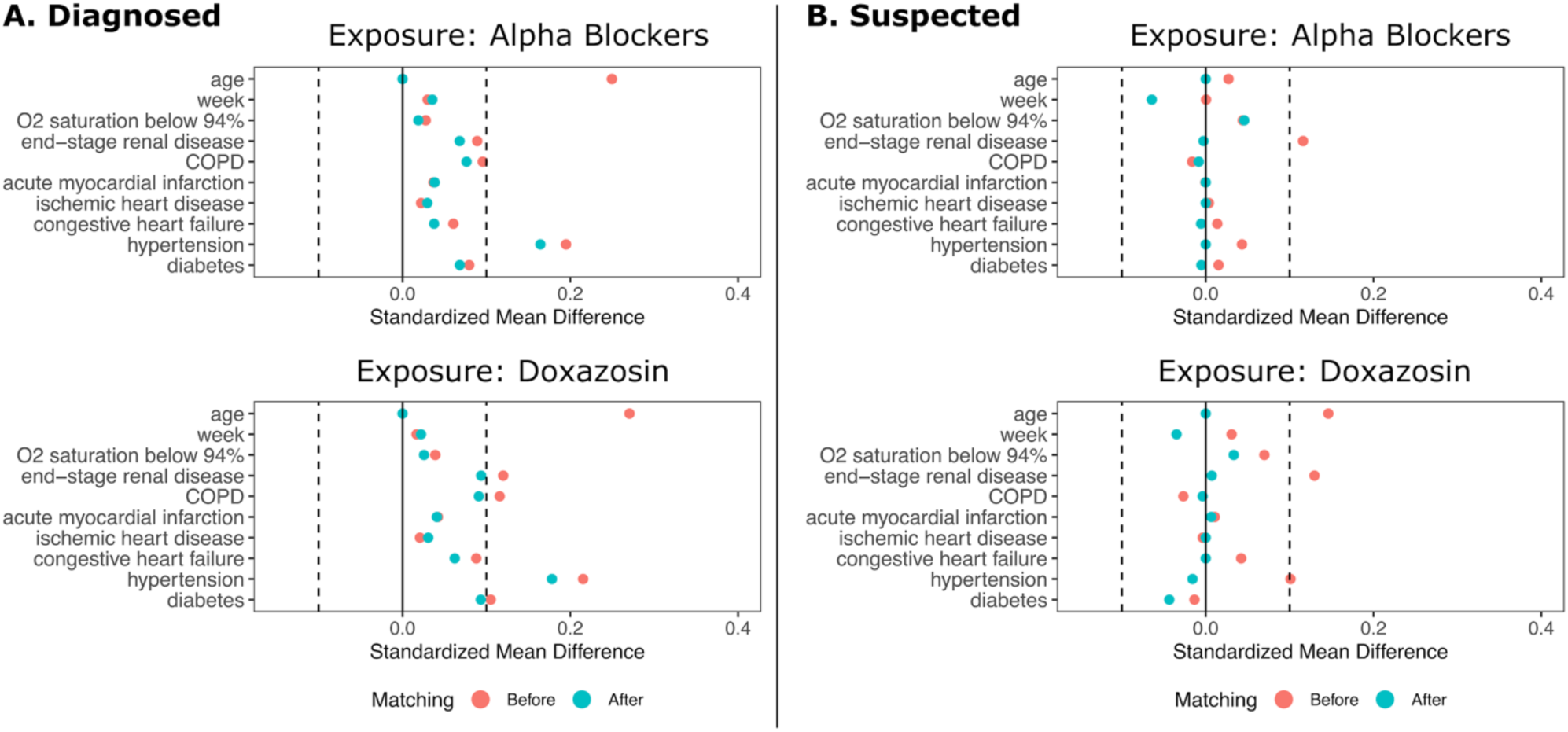
Standardized Mean Differences in Patient Characteristics Before and After Matching. The left panel shows the results for patients diagnosed with COVID-19. The right panel shows the results for patients diagnosed with COVID-19 plus suspected COVID-19 patients. Top panel show data for any α_1_-AR antagonists; bottom panel show data for doxazosin.

## 3 Results

### 3.1 Sample Characteristics

The sample contained 25,130 patients with COVID-19, with 5,600 patients taking any α_1_-AR antagonist at time of admission. Of those taking α_1_-AR antagonists, 73% of patients were on tamsulosin (n=4,078), 12% on terazosin (n=679), 10% on prazosin (n=581), 4% on doxazosin (n=215), 3% on alfuzosin (n=186), and less than 1% were on silodosin (n=5) (Figure 1). 177 patients had active prescriptions for more than one α_1_-AR antagonist at the time of admission. Demographic characteristics, medical comorbidities, and Charlson Comorbidity Index for patient groups prior to matching is shown in Table 1. The differences in sample characteristics after matching are summarized in Figure 3.

**Table 1.**
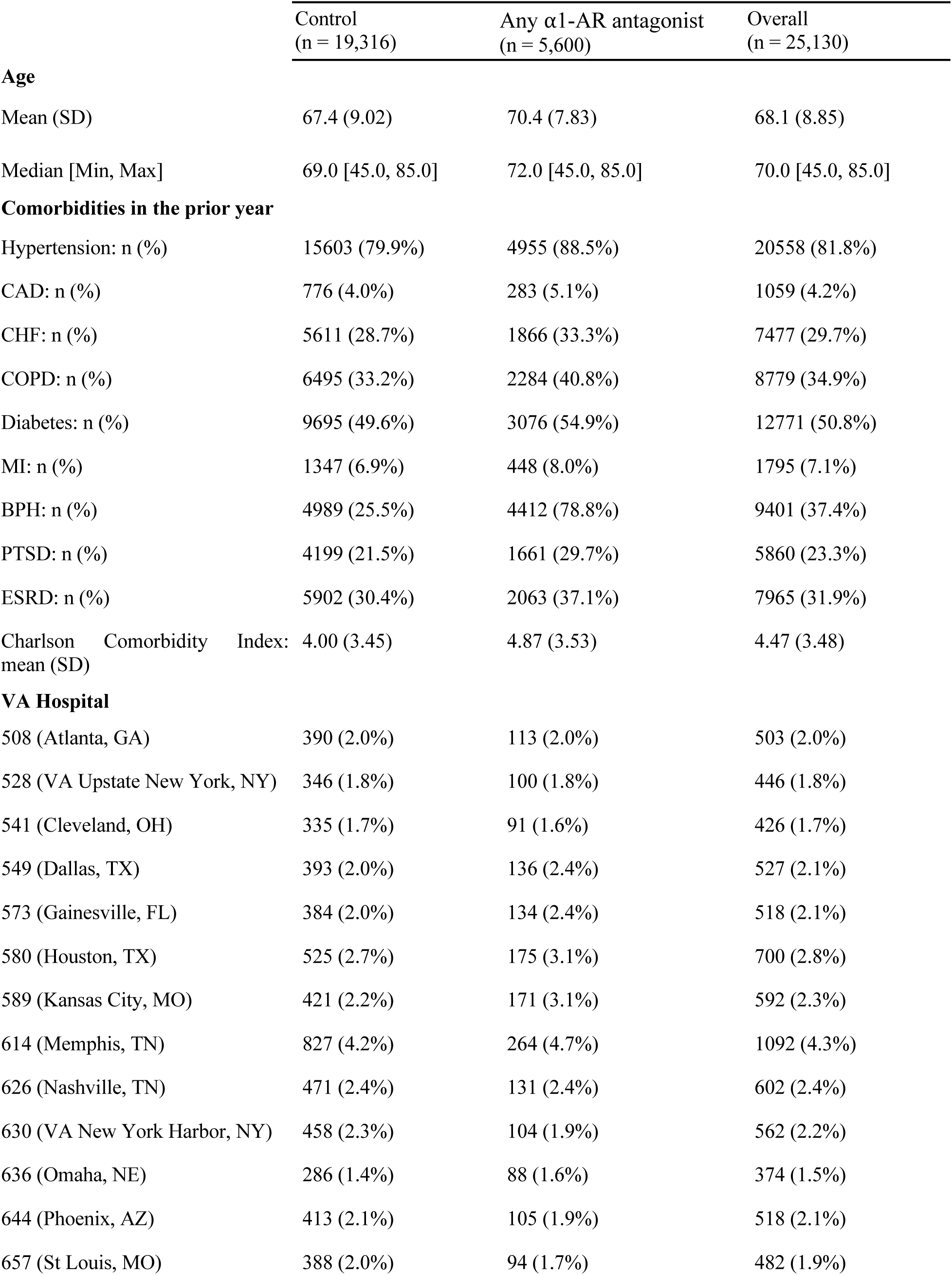
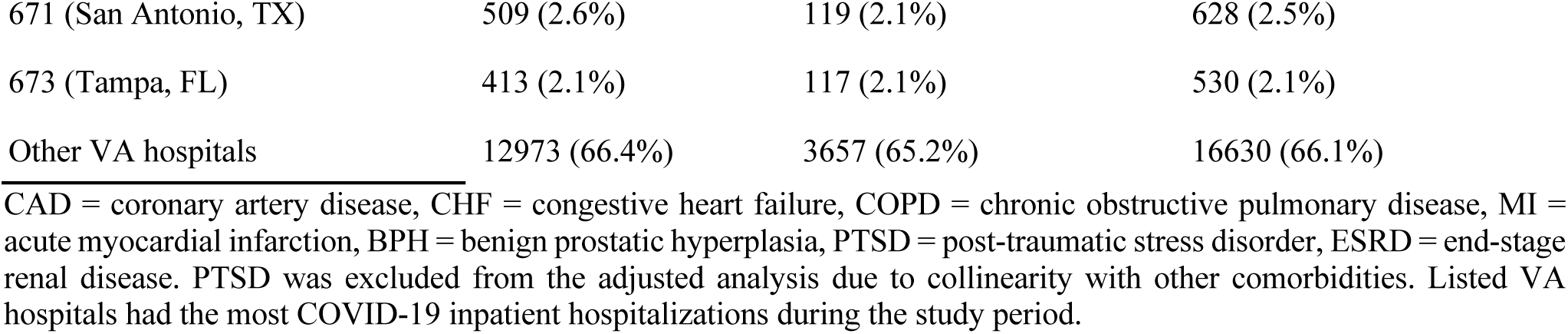
Patient and Sample Characteristics at Time of Admission.

| | Control<br>(n = 19,316) | Any α1-AR antagonist<br>(n = 5,600) | Overall<br>(n = 25,130) |
| --- | --- | --- | --- |
| **Age** |  |  |  |
| Mean (SD) | 67.4 (9.02) | 70.4 (7.83) | 68.1 (8.85) |
| Median [Min, Max] | 69.0 [45.0, 85.0] | 72.0 [45.0, 85.0] | 70.0 [45.0, 85.0] |
| **Comorbidities in the prior year** |  |  |  |
| Hypertension: n (%) | 15603 (79.9%) | 4955 (88.5%) | 20558 (81.8%) |
| CAD: n (%) | 776 (4.0%) | 283 (5.1%) | 1059 (4.2%) |
| CHF: n (%) | 5611 (28.7%) | 1866 (33.3%) | 7477 (29.7%) |
| COPD: n (%) | 6495 (33.2%) | 2284 (40.8%) | 8779 (34.9%) |
| Diabetes: n (%) | 9695 (49.6%) | 3076 (54.9%) | 12771 (50.8%) |
| MI: n (%) | 1347 (6.9%) | 448 (8.0%) | 1795 (7.1%) |
| BPH: n (%) | 4989 (25.5%) | 4412 (78.8%) | 9401 (37.4%) |
| PTSD: n (%) | 4199 (21.5%) | 1661 (29.7%) | 5860 (23.3%) |
| ESRD: n (%) | 5902 (30.4%) | 2063 (37.1%) | 7965 (31.9%) |
| Charlson Comorbidity Index: mean (SD) | 4.00 (3.45) | 4.87 (3.53) | 4.47 (3.48) |
| **VA Hospital** |  |  |  |
| 508 (Atlanta, GA) | 390 (2.0%) | 113 (2.0%) | 503 (2.0%) |
| 528 (VA Upstate New York, NY) | 346 (1.8%) | 100 (1.8%) | 446 (1.8%) |
| 541 (Cleveland, OH) | 335 (1.7%) | 91 (1.6%) | 426 (1.7%) |
| 549 (Dallas, TX) | 393 (2.0%) | 136 (2.4%) | 527 (2.1%) |
| 573 (Gainesville, FL) | 384 (2.0%) | 134 (2.4%) | 518 (2.1%) |
| 580 (Houston, TX) | 525 (2.7%) | 175 (3.1%) | 700 (2.8%) |
| 589 (Kansas City, MO) | 421 (2.2%) | 171 (3.1%) | 592 (2.3%) |
| 614 (Memphis, TN) | 827 (4.2%) | 264 (4.7%) | 1092 (4.3%) |
| 626 (Nashville, TN) | 471 (2.4%) | 131 (2.4%) | 602 (2.4%) |
| 630 (VA New York Harbor, NY) | 458 (2.3%) | 104 (1.9%) | 562 (2.2%) |
| 636 (Omaha, NE) | 286 (1.4%) | 88 (1.6%) | 374 (1.5%) |
| 644 (Phoenix, AZ) | 413 (2.1%) | 105 (1.9%) | 518 (2.1%) |
| 657 (St Louis, MO) | 388 (2.0%) | 94 (1.7%) | 482 (1.9%) |
| 671 (San Antonio, TX) | 509 (2.6%) | 119 (2.1%) | 628 (2.5%) |
| 673 (Tampa, FL) | 413 (2.1%) | 117 (2.1%) | 530 (2.1%) |
| Other VA hospitals | 12973 (66.4%) | 3657 (65.2%) | 16630 (66.1%) |

CAD = coronary artery disease, CHF = congestive heart failure, COPD = chronic obstructive pulmonary disease, MI = acute myocardial infarction, BPH = benign prostatic hyperplasia, PTSD = post-traumatic stress disorder, ESRD = end-stage renal disease. PTSD was excluded from the adjusted analysis due to collinearity with other comorbidities. Listed VA hospitals had the most COVID-19 inpatient hospitalizations during the study period.

### 3.2 Risk of In-Hospital and 28-Day Mortality

For all patients admitted to VA hospitals between February 20, 2020, and October 7, 2020, the overall in-hospital mortality rate was 2.5%. Among hospitalized patients with confirmed COVID-19 (8.9% of all admissions), in-hospital mortality was 6% overall and 5.5% in our sample. Patients with confirmed COVID-19 taking any α_1_-AR antagonist, compared to non-users, had an 18% relative risk reduction for death during their hospitalization (243/5,309=4.6% in matched treatment group vs. 984/17,538=5.6% in matched control group, p ≤ 0.001, Figure 2) and a 17% relative risk reduction for death within 28 days from the date of admission (331/5,309=6.2% in matched treatment group vs. 1,318/17,538=7.5% in matched control group, p ≤ 0.001, Figure 2).

The top panel of Figure 2 shows the unadjusted, propensity score adjusted, and matched odds ratios among patients diagnosed with COVID-19 (n=25,130). The bottom panel expands the denominator to also include patients with suspected COVID-19 (n=32,016). The dark green odds ratios in Figure 2 represent all α_1_-AR antagonists, while the lighter green represent doxazosin. Results were similar for the suspected COVID-19 sample. Patients taking any α_1_-AR antagonists, compared to non-users, had an 20% relative risk reduction for death (p ≤ 0.001) in this cohort (Figure 2).

The use of doxazosin, a nonselective α_1_-AR antagonist targeting all three α_1_-AR subtypes, resulted in a 74% relative risk reduction for death in hospitalized patients with COVID-19 during the index admission (2/155=1.3% in matched treatment group vs. 39/775=5.0% in matched control group, odds ratio for death 0.23; p=0.028, Figure 2). Use of tamsulosin, the most commonly prescribed α_1_-AR antagonist in this cohort with selectivity for α_1A_- and α_1D_-ARs, was associated with a 18% relative risk reduction for death during the inpatient stay (odds ratio for death 0.77; p=0.002, Supplementary Figure 2). Even though COVID-19 has affected different parts of the United States at different times, we found no evidence that these results were driven by any particular time period or location (Supplementary Figures 3 and 4).

## 4 Discussion

In this retrospective analysis of patients with COVID-19, we found a significant negative association between the use of α_1_-AR antagonists and in-hospital or 28-day mortality. These results are consistent with findings from a recent retrospective study of >300,000 patients hospitalized with pneumonia or ARDS unrelated to SARS-CoV-2 infection that identified a significant risk reduction for the progression to mechanical ventilation and death in individuals who were receiving any α_1_-AR antagonists as compared to non-users (5), suggesting that the benefits of α_1_-AR inhibition for mortality may be independent of etiology in patients with lower respiratory tract infection or inflammation.

Interestingly, we found much larger effect sizes in reducing mortality for patients treated with doxazosin, an antagonist on all three α_1_-AR subtypes (α_1A_-, α_1D_-, and α_1B_-AR), than for a pooled population of patients treated with any α_1_-AR antagonist in whom tamsulosin was the most common drug (72%). This was similarly true for patients treated exclusively with tamsulosin, a “uroselective” α_1_-AR antagonist on α_1A_- and α_1D_-ARs without clinically relevant inhibition of α_1B_-ARs expressed by immune cells and the peripheral vasculature (20). In patients with test-confirmed COVID-19, baseline use of doxazosin was associated with significantly reduced in-hospital and 28-day mortality compared to controls (odds ratio for death during admission 0.19 in adjusted cohort; odds ratio and relative risk reduction for death 0.23 and 74% in matched cohort, respectively). Baseline use of tamsulosin in patients with confirmed COVID-19, by comparison, was associated with significant, but less pronounced reductions in mortality. A similar trend was previously observed in patients with pneumonia in whom use of doxazosin was associated with lower risk of mechanical ventilation and death than tamsulosin (13). These observed differences in effect size are biologically plausible and may reflect the distinct pharmacological selectivity of doxazosin and tamsulosin for α_1_-AR subtypes.

Immune cells can induce expression of all three α_1_-AR subtypes (i.e., α_1A_-, α_1D_-, and α_1B_-ARs (21), and catecholamine signaling through these individual receptors may be highly redundant (12). As such, α_1_-AR antagonists acting on all three receptor subtypes (i.e., doxazosin, prazosin, alfuzosin, terazosin) may be required to effectively interrupt autocrine and paracrine catecholamine signaling in monocytes and other immune cells that enhance inflammatory injury (14,20). Indeed, preclinical data suggests that nonselective α_1_-AR antagonists are effective in preventing hyperinflammation and death in animal models of cytokine storm syndrome (11). The markedly improved survival in patients on doxazosin as compared to tamsulosin or any α_1_-AR antagonist (a cohort highly enriched in tamsulosin use) may therefore be consistent with a redundancy in catecholamine signaling pathways which are globally inhibited by doxazosin, whereas tamsulosin allows for continued signaling through the α_1B_-AR. These findings have practical implications for the selection of α_1_-AR antagonists for the prevention of inflammatory injury and suggest that the immunomodulatory benefits may not be uncoupled from inhibition of α_1B_-ARs expressed on the peripheral vasculature.

Additional studies have explored the efficacy of α_1_-AR blockade in the prevention of inflammatory and autoimmune injury. In a model of encephalitis, early α_1_-AR inhibition reversed neutrophil infiltration in lungs and prevented hemorrhagic pulmonary edema (22). The non-selective α_1_-AR antagonist prazosin has been shown to ameliorate experimental autoimmune encephalomyelitis (23). In a preclinical model of ischemia-reperfusion injury, prazosin administration resulted in decreased expression of IL-6, TNF-α, IL-10, and IL-1, and prevented mortality (24). Finally, α_1_-AR antagonism has been shown to block cytokine production in human peripheral blood mononuclear cells from patients with juvenile polyarticular arthritis, and treatment with doxazosin abrogated catecholamine-augmented secretion of IL-6 (25). These studies suggest a role of catecholamine-associated augmentation of injurious cytokine responses beyond cytokine release syndrome and acute lung infection and highlights the potential of α_1_-AR antagonists across various inflammatory diseases.

One concern with observational analysis is confounding by indication, especially if medications given during a hospital stay are correlated with disease severity. To avoid confounding by indication, this analysis examined the use of α_1_-AR antagonists prior to index hospitalization. This class of medications is primarily used to manage chronic diseases such as arterial hypertension, PTSD, or BPH. As such, prescribing practices would not be biased by the severity of COVID-19. In addition, our results were not driven by a specific location or time period.

This study has important strengths and weaknesses. We have focused on mortality as a definitive clinical outcome, thereby avoiding process measures, such as use of mechanical ventilators or admission to an ICU, that are subject to local and individual practice patterns and would be biased if clinicians or hospitals changed their practices in unobserved ways. Another strength is our use of information prior to the COVID-19 admission for risk adjustment. One limitation in this study was the exclusion of women which was required due to limitations in samples size since α_1_-AR antagonists are most commonly used to treat benign prostatic hyperplasia and 90% of patients in the VA system are men (26). A second limitation, best addressed in prospective clinical trials, was our inability to examine dose effects given our sample size.

Our results suggest that inhibition of catecholamine signaling with doxazosin (and other α_1_-AR antagonists) may reduce in-hospital and 28-day mortality in patients with COVID-19 and highlight the need for randomized placebo-controlled clinical trials to examine the efficacy of α_1_-AR antagonists for improving survival and preventing adverse outcomes from COVID-19. Importantly, α_1_-AR antagonists are inexpensive, administered orally, do not require refrigeration, and have a well-established safety profile. Thus, if trials confirm these results, α_1_-AR antagonists could be widely deployed to reduce mortality from inflammatory injury. Importantly, α_1_-AR antagonists are immunomodulatory, but not immunosuppressive drugs. Long-term use of doxazosin does not appear to be associated with the development of opportunistic infection in human studies (27). Indeed, some studies suggest an overall decreased risk of urinary tract infection compared to placebo as may be expected based on its effect on dynamic prostate and bladder function (28). The absence of serious infectious complications may be explained by the unique mechanism of action of α_1_-AR antagonists compared to immunosuppressive drugs currently employed in the treatment of severe COVID-19 (e.g., dexamethasone, baricitinib, tocilizumab) which confer an increased risk of opportunistic infection.

In summary, patients hospitalized with COVID-19 had lower odds of in-hospital and 28-day death if they had an active prescription for any α_1_-AR antagonist (tamsulosin, silodosin, prazosin, terazosin, doxazosin, or alfuzosin) at the time of admission. Among different α_1_-AR antagonists, doxazosin was associated with a 74% relative risk reduction for death, while tamsulosin had a more modest 18% relative risk reduction for death. A clinical trial testing the efficacy and safety of α_1_-AR antagonists such as doxazosin to prevent hyperinflammation and reduce mortality in COVID-19 would appear warranted.

## Data Availability

The data used in this study includes PHI held by the US Veterans Health Administration and is not publicly available.

## 6 Author Contributions

JV, BV, CB, and MK conceived of the presented idea. LR, LG, AK, MP, and RX conducted statistical analyses and results presentation. AK, MP, RX, ZS, and SA developed the methodology. LR, MK, JV, TW wrote the manuscript with input from all authors. All authors reviewed the final manuscript.

## 8 Acknowledgments

Any opinion, findings, and conclusions or recommendations expressed in this material are those of the authors and do not necessarily reflect the views of the US Department of Veterans Affairs or the National Science Foundation.

This project was sponsored by the VA Office of Reporting, Analytics, Performance, Improvement and Deployment, and the VA Center for Medication Safety, and approved as quality improvement by the Stanford University Institutional Review Board.

---

**Supplementary Figure 1.**
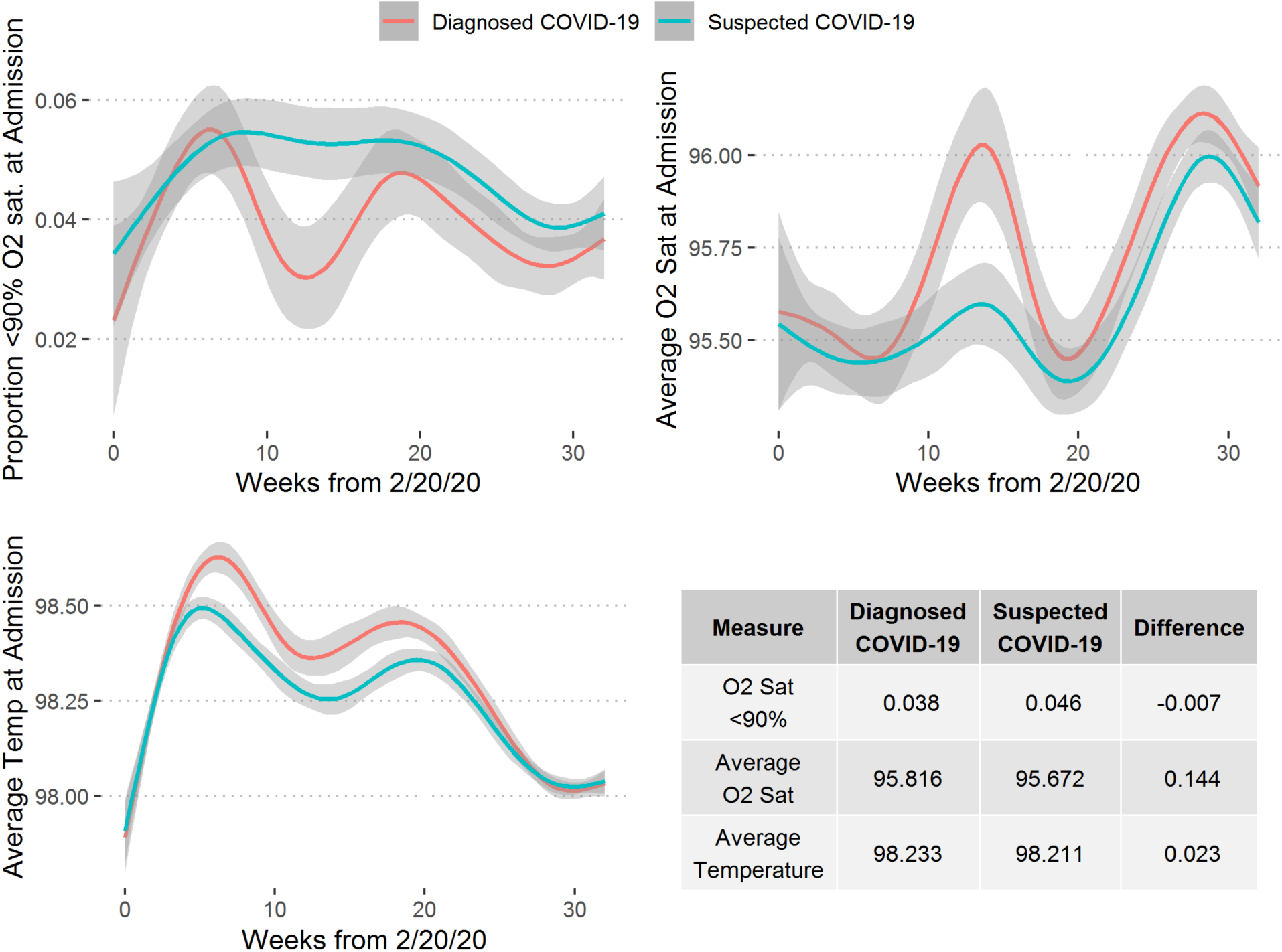
Vital Signs at Time of Admission. The diagrams show vital signs for patients diagnosed with COVID-19 (red line) and an expanded cohort of patients with suspected COVID-19 (blue line). Smoothed lines are from a LOESS model with 95% confidence intervals shown (gray ribbons).

**Supplementary Figure 2.**
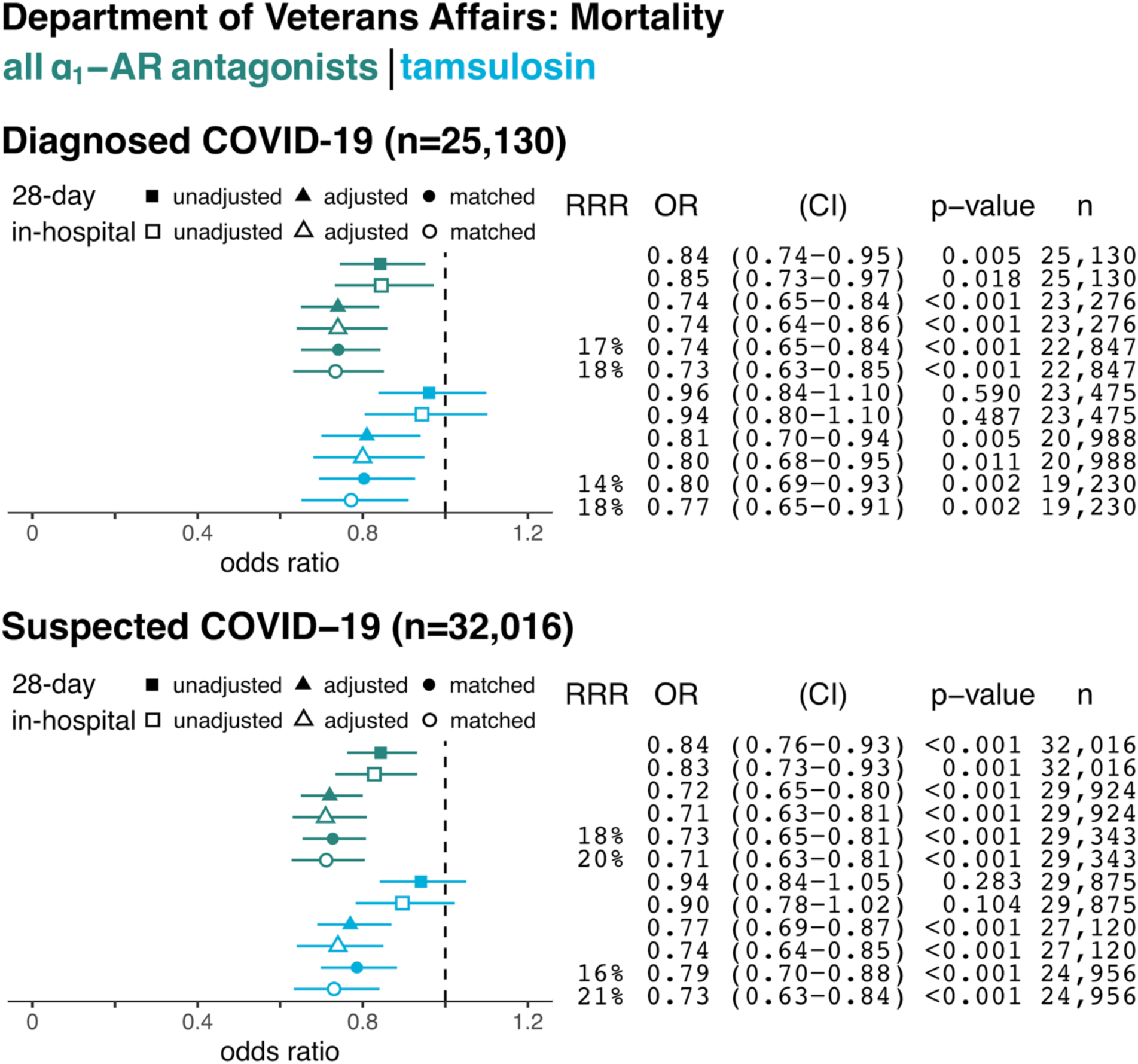
In-hospital and 28-Day Mortality by Use of Tamsulosin at Time of Hospital Admission with COVID-19. Data are shown for hospitalized patients diagnosed with confirmed COVID-19 (top panel) and with confirmed plus suspected COVID-19 (bottom panel). Forest plots showing odds ratios (OR) of in-hospital mortality based on prior use of any alpha-1 adrenergic receptor antagonists (dark green) or tamsulosin (light blue) in each panel. Relative risk reduction (RRR), odds ratios (ORs) for death, 95% confidence intervals (CI), and p-values (for unadjusted, adjusted, and matched models), and sample size (n) for each analysis are shown on the right.

**Supplementary Figure 3.**
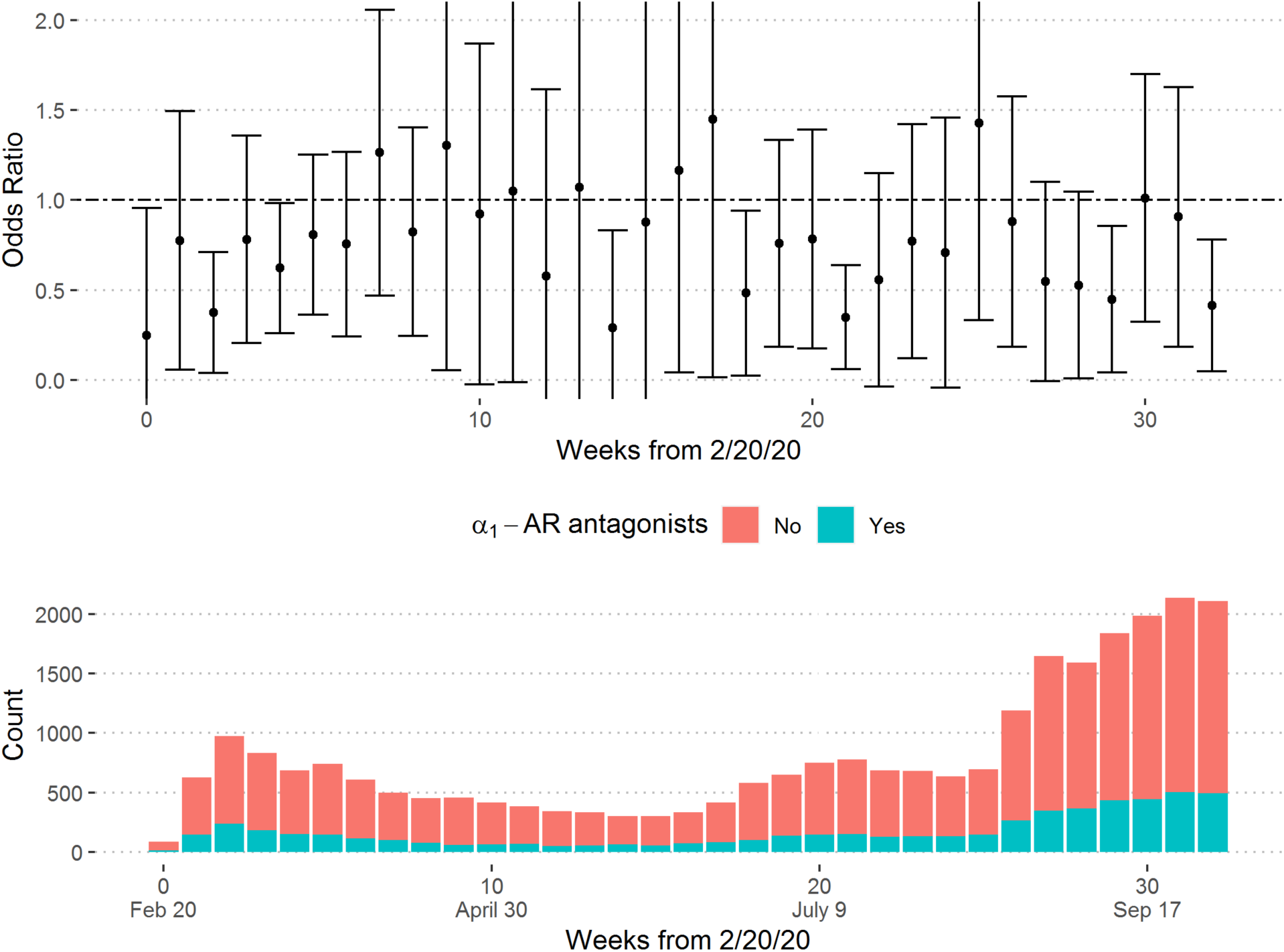
Adjusted Odds of In-hospital Mortality and Use of α_**1**_-AR Antagonists by Week. Top panel shows adjusted odds ratios of in-hospital mortality and use of α_1_-AR antagonists by week of admission. Top panel truncated between 0 and 2 to aid visualization. Bottom panel shows number of new admissions by week and use of α_1_-AR antagonists (bottom).

**Supplementary Figure 4.**
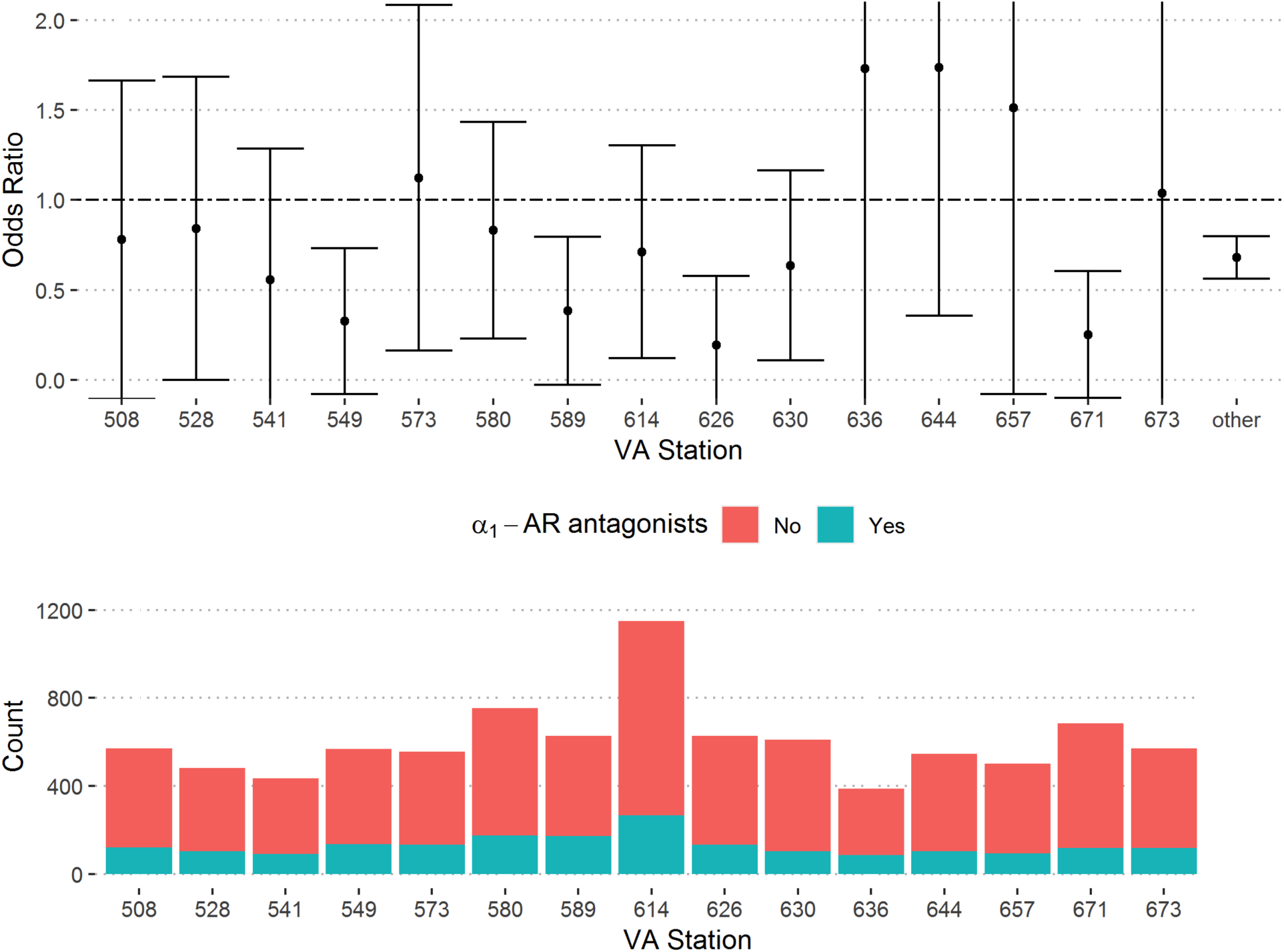
Adjusted Odds of In-hospital Mortality and Use of α_**1**_-AR Antagonists by VA Station. Top panel shows adjusted odds ratios of in-hospital mortality in patients taking α_1_-AR antagonists by VA station. Top panel truncated between 0 and 2 to aid visualization. Bottom panel shows number of new admissions and use of α_1_-AR antagonists by VA station (bottom). For other VA stations, the number of admissions of patients not using α_1_-AR antagonists was 7,645 and number of admissions of patients using α_1_-AR antagonists was 1,845. VA stations shown: 508 = Atlanta, 549 = Dallas, 573 = Gainesville, 580 = Houston, 589 = Kansas City, 614 = Memphis, 630 = New York Harbor, 644 = Phoenix, 671 = San Antonio, 673 = Tampa.

